# Cardiometabolic Health in Asian American Children

**DOI:** 10.1101/2023.11.11.23298417

**Authors:** Julian Sethna, Kristal Wong, Kevin Meyers

## Abstract

**Background:** The aim was to compare cardiometabolic health between Asian American children and Non-Hispanic White (NHW) children as well as to compare cardiometabolic health among Asian American children by birthplace.

**Methods:** Children aged 6-17 years enrolled in the National Health and Nutrition Examination Survey (NHANES) from 2011-2018 who self-identified as non-Hispanic Asian and NHW were included. Among Asian Americans, place of birth was defined as foreign-born vs United States (US)-born. Regression models were adjusted for age, sex, household income, food insecurity, passive smoke exposure, and body mass index (BMI) z-score.

**Results:** Among 3369 children, 8.4% identified as Asian American (age 11.7 years) and 91.6% identified as NHW (age 11.7 years). Compared to NHW children, Asian American children had significantly lower BMI z-scores and odds of obesity. Asian American children had higher HOMA-IR and uric acid, and greater odds of dyslipidemia, microalbuminuria and glomerular hyperfiltration compared to NHW children. Among Asian Americans, 30.5% were foreign-born. Compared to foreign-born Asian American children, US-born Asian American children had significantly higher non-HDL, triglycerides, HOMA-IR and uric acid, lower HDL, and lower odds of hyperfiltration. There were no differences in blood pressure by racial group or place of birth.

**Conclusions:** Although Asian American children have lower odds of obesity, they have significantly worse glucose intolerance, higher serum uric acid levels, more dyslipidemia and more microalbuminuria compared to NHW children. US-born Asian American children have worse cardiometabolic health profiles compared to foreign-born Asian Americans.

## Introduction

The United States (US) Census estimates the population of Asian Americans in 2020 to be approximately 24 million [1]. Asian Americans have emerged as the fastest growing racial/ethnic group in the country, increasing 81% between 2000 and 2019. By 2060, the Asian American population is projected to reach 35 million [2]. This period of significant population growth provides a critical opportunity to study the health outcomes and health disparities that may be unique to Asian Americans.

Recent literature has highlighted differences in the cardiometabolic profiles between Asian Americans and Non-Hispanic Whites (NHWs) in the adult population. Studies have found that Asian Americans had overall better cardiovascular health and less cardiovascular disease when compared to NHWs [3,4]. In contrast, other studies have reported that Asian Americans had a higher prevalence of diabetes and obesity compared to NHWs [5]. Among Asian American subgroups, further disparities exist; Indians had the highest prevalence of diabetes, while Filipinos had the highest prevalence of hyperlipidemia, hypertension, and obesity. Chinese participants experienced the lowest odds of cardiovascular disease risk factors [4]. Additional research has found differences in cardiometabolic outcomes when comparing place of birth amongst Asian Americans [5].

Despite the growing recognition of health disparities within the Asian American adult population, medical understanding of cardiometabolic health among Asian American children remains limited. Therefore, the purpose of this study was to compare the cardiometabolic health profiles of Asian American children and NHW children, including measures of adiposity, blood pressure, lipids, glucose intolerance, kidney markers and uric acid. Additionally, this study sought to explore potential differences in cardiometabolic health among Asian American children by birthplace. It was hypothesized that Asian American children would exhibit lower rates of obesity and better cardiometabolic health profiles compared to NHW children and that among Asian American children, those who were foreign born would demonstrate better cardiometabolic health profiles compared to those who were US born.

## Methods

### Study Population

This study was a cross-sectional analysis of the National Health and Nutrition Examination Survey (NHANES) conducted between 2011 and 2018. NHANES is a nationally representative survey used to assess the health and nutrition of both adults and children in the US [6].

Participants completed interviews, medical exams, and 24-hour dietary recalls. The criteria for inclusion were children ages 6-17 who self-identified as Asian American or Non-Hispanic White. Those who were pregnant, born prematurely, or had a history of major health problems were excluded. Prior to participating in NHANES, participants/guardians provided informed consent/assent to NHANES and the study was reviewed by the National Center for Health Statistics and Ethics Review Board [7].

### Race/Ethnicity Exposure

The primary exposure was self-identified race, specifically comparing Asian American children to NHW children. The Asian American category included participants who self-reported as non-Hispanic and non-Black Asian of single race or in combination with another race and excluded Black individuals [3,8]. The second exposure, participant place of birth, compared US born to foreign born birthplace within participants in the Asian American group [6].

### Cardiometabolic Outcomes

The primary outcomes of interest were obesity and blood pressure. Body mass index (BMI) z-scores were calculated. Obesity was defined as BMI greater than the 95^th^ percentile [9]. Systolic and diastolic blood pressures were measured via auscultation [10]. Up to four measurements were obtained and averaged for analysis. For children aged 6-12 years, blood pressure was indexed to the 95^th^ percentile for age, sex, and height. Hypertensive blood pressure was defined as systolic or diastolic blood pressure index ≥1 [11]. For adolescents ≥13 years, hypertensive blood pressure was defined as systolic blood pressure ≥130 mmHg or diastolic blood pressure ≥80 mmHg [11].

Secondary cardiometabolic outcome measures of interest included lipids, uric acid, insulin resistance, glomerular hyperfiltration, and microalbuminuria. Dyslipidemia was defined as triglycerides ≥130 mg/dL, HDL cholesterol <40 mg/dL, or LDL cholesterol ≥160 mg/dL [12]. Homeostatic Model Assessment of Insulin Resistance (HOMA-IR) was calculated using the formula: (Insulin x Glucose)/22.5. Glomerular hyperfiltration was defined by estimated glomerular filtration rate (eGFR) >140 ml/min/1.73m² using the new Schwartz formula [13]. Microalbuminuria was defined as urine albumin:creatinine >30 mg/g.

### Dietary Factors and Confounding Variables

From the 24-hour dietary recall, NHANES used the United States Department of Agriculture (USDA) Food and Nutrient Database for Dietary Studies to estimate the amount of food energy and nutritional components consumed [10]. Consumption of dietary energy, sugar, fat, protein, potassium, magnesium, cholesterol, dietary fiber, and carbohydrates were recorded from the Day 1 dietary recall.

Potential confounding variables of age, sex, household income, food insecurity, and passive smoke exposure were collected. Food insecurity was defined as those who answered low or very low household food security on the NHANES questionnaire.

### Statistical Analysis

All complex sample statistics were performed using SPSS version 28 (IBM Inc). P values <0.05 were considered statistically significant. Weighting adjustments of the data accounted for the 4 two-year cycles of NHANES data. Descriptive statistics included means with standard error (SE) and frequencies with percentages of each population statistic. T-test and Chi square were used to compare demographic, clinical and dietary variables between Asian American and NHW children. To further examine differences in cardiometabolic health between the two groups, linear and logistic regression models were adjusted for the following variables chosen *a priori*: age, sex, household income, food insecurity, passive smoke exposure, and BMI z-score. Forest plots were used to graphically present regression data. T-test, Chi square, and regression models were also applied to the Asian American group to determine differences in cardiometabolic health between US born and foreign-born birthplace.

## Results

A total of 3,369 children met eligibility criteria and were included in analysis. The study population included 8.4% self-identified Asian Americans, with a mean age of 11.7 years and sex distribution of 50.4% male. Among Asian children, 30.5% were foreign born. Self-identified NHW participants encompassed 91.6% of the study population, with a mean age of 11.7 years and sex distribution of 50.8% male. Demographic and clinical characteristics comparing Asian American and NHW children are presented in Table 1. Asian American children had significantly lower BMI z-score (p<0.001) and higher uric acid level (p=0.01) compared to NHW children. Although obesity was more prevalent among NHW (10.1%) compared to Asian Americans (4.2%), the difference was not statistically significant. There were no other significant differences in blood pressure or other cardiometabolic markers between the two groups.

**Table 1.** Demographic and Clinical Characteristics of Asian American and Non-Hispanic White Children in NHANES

| N=3369 | Asian American<br>N= 949 | Non-Hispanic White<br>N= 2420 | P-Value |
| --- | --- | --- | --- |
| Mean (SE) or % | 8.4% | 91.6% |  |
| Age | 11.7 (0.9) | 11.7 (0.9) | 0.89 |
| Male | 50.4% | 50.8% | 0.87 |
| Household income <45K | 22.2% | 25.5% | 0.19 |
| Food Insecurity | 6.2% | 6.7% | 0.29 |
| Passive smoke exposure | 10.2% | 20.5% | 0.07 |
| BMI Z-score | 0.23 (0.07) | 0.67 (0.05) | <0.001 |
| Obesity | 4.2% | 10.1% | 0.08 |
| SBP Index | 0.79 (0.01) | 0.81 (0.01) | 0.31 |
| DBP Index | 0.7 (0.01) | 0.71 (0.01) | 0.42 |
| Hypertensive BP | 2% | 2% | 0.98 |
| Dyslipidemia | 19% | 20.3% | 0.75 |
| HOMA-IR | 2.88 (0.21) | 2.78 (0.15) | 0.19 |
| Uric Acid | 5.18 (0.1) | 5.02 (0.06) | 0.01 |
| eGFR | 100.99 (3.21) | 100.68 (3.21) | 0.67 |
| Hyperfiltration | 6.4% | 4.2% | 0.26 |
| Microalbuminuria | 14% | 10.8% | 0.31 |
| SE- standard error, BMI- body mass index, SBP- systolic blood pressure, DBP- diastolic blood pressure, BP- blood pressure, HOMA-IR- Homeostatic Model Assessment of Insulin Resistance, eGFR- glomerular filtration rate |  |  |  |

### Dietary Factors

Dietary variables by racial group and birthplace among Asian Americans are presented in Table 2. NHW children consumed a greater amount of sugar compared to Asian American children (p = 0.003). There were no significant differences in consumption of energy, sodium, potassium, fat, magnesium, carbohydrates, cholesterol, or dietary fiber between the two groups. Additionally, there were no significant differences in consumption of any dietary variables between Asian Americans who were US born and those who were foreign born.

**Table 2.** Dietary Variables by Race and Birthplace among Asian American Children in NHANES

| Dietary Variables | Asian | Non-Hispanic White | P value | US Born | Foreign Born | P value |
| --- | --- | --- | --- | --- | --- | --- |
| Energy, kcal | 1970.3 (88.9) | 2059.3 (55.7) | 0.4 | 1987.9 (57.6) | 1940.3 (100.4) | 0.56 |
| Sodium, mg | 3325.3 (134.4) | 3353.3 (106.5) | 0.87 | 3407.6 (100.4) | 3179.7 (147.5) | 0.13 |
| Sugar, gm | 103.4(6.3) | 126.2(3.5) | 0.003 | 104.6 (5.2) | 101.3 (9.7) | 0.72 |
| Potassium, mg | 2323.0 (112.8) | 2215.8 (87.5) | 0.46 | 2297.7 (51.0) | 2368 (178.7) | 0.66 |
| Fat, gm | 74.5 (4,.2) | 80 (2.8) | 0.27 | 75.0 (2.2) | 73.6 (4.3) | 0.70 |
| Protein,gm | 76.8 (4.1) | 74.8 (3.2) | 0.71 | 78.2 (3.2) | 74.4 (5.2) | 0.34 |
| Magnesium, mg | 263.6 (11.6) | 248.2 (9.1) | 0.28 | 257.3 (7.0) | 274.6 (17.5) | 0.35 |
| Carbohydrates, gm | 253.7 (11.0) | 264.9 (6.8) | 0.4 | 2.1 (0.1) | 3.1 (0.5) | 0.13 |
| Cholesterol, mg | 247.4 (16.8) | 257 (15.3) | 0.67 | - | - | - |
| Dietary fiber, gm | 15.9 (.77) | 14.1 (.49) | 0.07 | - | - | - |

### Cardiometabolic Variables: Asian vs Non-Hispanic White

In adjusted regression analysis, the Asian American group (reference NHW) was associated with lower BMI z-score (b = -0.36, 95% CI -0.57, -0.15 p=0.001) and 0.48 lower odds of obesity. There were no associations between racial group and blood pressure. Asian American children had higher HOMA-IR (b =0.98, 95% CI 0.38, 1.59 p=0.002) and higher uric acid (b = 0.33, 95% CI 0.02, 0.64 p=0.04) as well as 2.2 higher odds of dyslipidemia, 2.1 higher odds of microalbuminuria, and 3.5 higher odds of hyperfiltration compared to NHW children (Table 3 and Figure 1).

**Table 3.** Adjusted Regression Analysis by Race and Birthplace among Asian American Children in NHANES*

|  |  | Asian American vs<br>Non-Hispanic White (ref) | US-born vs Foreign-born (ref) |
| --- | --- | --- | --- |
| $\beta$ (95% CI) | | | |
| BMI z-score | Crude<br>Adjusted | -0.48 (-0.64, -0.25) p <0.001<br>-0.36 (-0.57, -0.15) p=0.001 | -0.33 (-0.71, 0.05) p= 0.08<br>0.18 (-0.07, 0.42) p=0.14 |
| SBP Index | Crude<br>Adjusted | -0.01 (-0.04, 0.02) p= 0.45<br>-0.02 (-0.06, 0.01) p=0.12 | -0.06 (-0.11, -0.01) p= 0.01<br>0.001 (-0.05, 0.05) p=0.96 |
| DBP Index | Crude<br>Adjusted | -0.01 (-0.06, 0.04) p= 0.63<br>-0.04 (-0.15, 0.000006) p=0.05 | -0.03 (-0.07, 0.02) p= 0.21<br>0.03 (-0.002, 0.07) p=0.06 |
| Non-HDL | Crude<br>Adjusted | 0.65 (-6.16, 4.46) p= 0.85<br>7.44 (-0.49, 15.38) p=0.07 | 5.31 (-8.48, 19.10) p= 0.43<br>26.55 (15.08, 38.03) p<0.001 |
| Triglycerides | Crude<br>Adjusted | 2.19 (-7.08, 11.45) p= 0.64<br>110.42 (9.09, 211.76) p=0.27 | 7.3 (-12.70, 27.35) p= 0.45<br>34.88 (12.72, 57.04) p=0.006 |
| HDL | Crude<br>Adjusted | 2.81 (0.92, 4.70) p= 0.004<br>-1.21 (-3.85, 1.44) p=0.36 | -1.25 (-4.63, 2.14) p =0.45<br>-4.00 (-6.88, -1.13) p=0.01 |
| LDL | Crude<br>Adjusted | 0.03 (-6.00, 6.07) p= 0.99<br>5.23 (-0.45, 10.92) p=0.07 | 3.11 (-7.76, 13.99) p= 0.56<br>19.82 (5.85, 33.78) p=0.01 |
| HOMA-IR | Crude<br>Adjusted | 0.01 (-0.42, 0.62) p= 0.70<br>0.98 (0.38, 1.59) p=0.002 | 0.252 (-0.29, 0.79) p= 0.34<br>0.56 (0.04, 1.09) p=0.03 |
| Urine albumin creatinine | Crude<br>Adjusted | 3.60 (-5.73, 12.93) p= 0.44<br>-3.61 (-15.43, 8.22) p = 0.54 | -0.02 (-12.78, 12.74) p= 0.99<br>-14.55 (-30.56, 1.87) p=0.08 |
| eGFR | Crude<br>Adjusted | 0.30 (-6.62, 7.23) p= 0.93<br>4.4 (-0.96, 9.76) p=0.1 | -7.75 (-12.67, -2.82) p= 0.004<br>-11.97 (-18.90, -5.04) p=0.004 |
| Uric Acid | Crude<br>Adjusted | 0.16 (-0.11, 0.43) p= 0.25<br>0.33 (0.02, 0.64) p=0.04 | -0.07 (-0.47, 0.34) p= 0.73<br>0.63 (0.38, 0.88) p<0.001 |
| Odds Ratio (95% CI) |  |  |  |
| Obesity | Crude<br>Adjusted | 0.39 (0.13, 1.16) p= 0.09<br>0.48 (0.28, 0.84) p=0.01 | 0.46 (0.05, 3.93) p= 0.46<br>2.03 (0.41, 10.12) p=0.36 |
| Hypertensive BP | Crude<br>Adjusted | 1.02 (0.26, 4.07) p= 0.98<br>1.28 (0.72, 2.69) p=0.40 | 2.67 (0.28, 25.40) p=0.37<br>0.41 (0.11, 0.15) p=0.17 |
| Dyslipidemia | Crude<br>Adjusted | 0.93 (0.57, 1.50) p= 0.75<br>2.21 (1.05, 4.64) p=0.04 | 0.86 (0.36, 2.05) p= 0.72<br>0.99 (0.02, 61.55) p=0.99 |
| Microalbuminuria | Crude<br>Adjusted | 1.34 (0.75, 2.39) p= 0.31<br>2.11 (1.31, 3.42) p=0.003 | 1.50 (0.62, 3.64) p= 0.35<br>2.81 (0.46, 17.31) p=0.23 |
| Hyperfiltration | Crude<br>Adjusted | 1.56 (0.71, 3.43) p= 0.26<br>3.51 (1.16, 10.63) p=0.03 | 0.94 (0.40, 2.22) p= 0.89<br>0.001 (0.00009, 0.004) p<0.001 |
| *Models adjusted for age, sex, household income, food insecurity, passive smoke exposure and BMI z-score<br>BMI- body mass index, SBP- systolic blood pressure, DBP- diastolic blood pressure, HDL- high density lipoprotein, LDL- low density lipoprotein, BP- blood pressure, HOMA-IR- Homeostatic Model Assessment of Insulin Resistance, eGFR- glomerular filtration rate |  |  |  |

**Figure 1:**
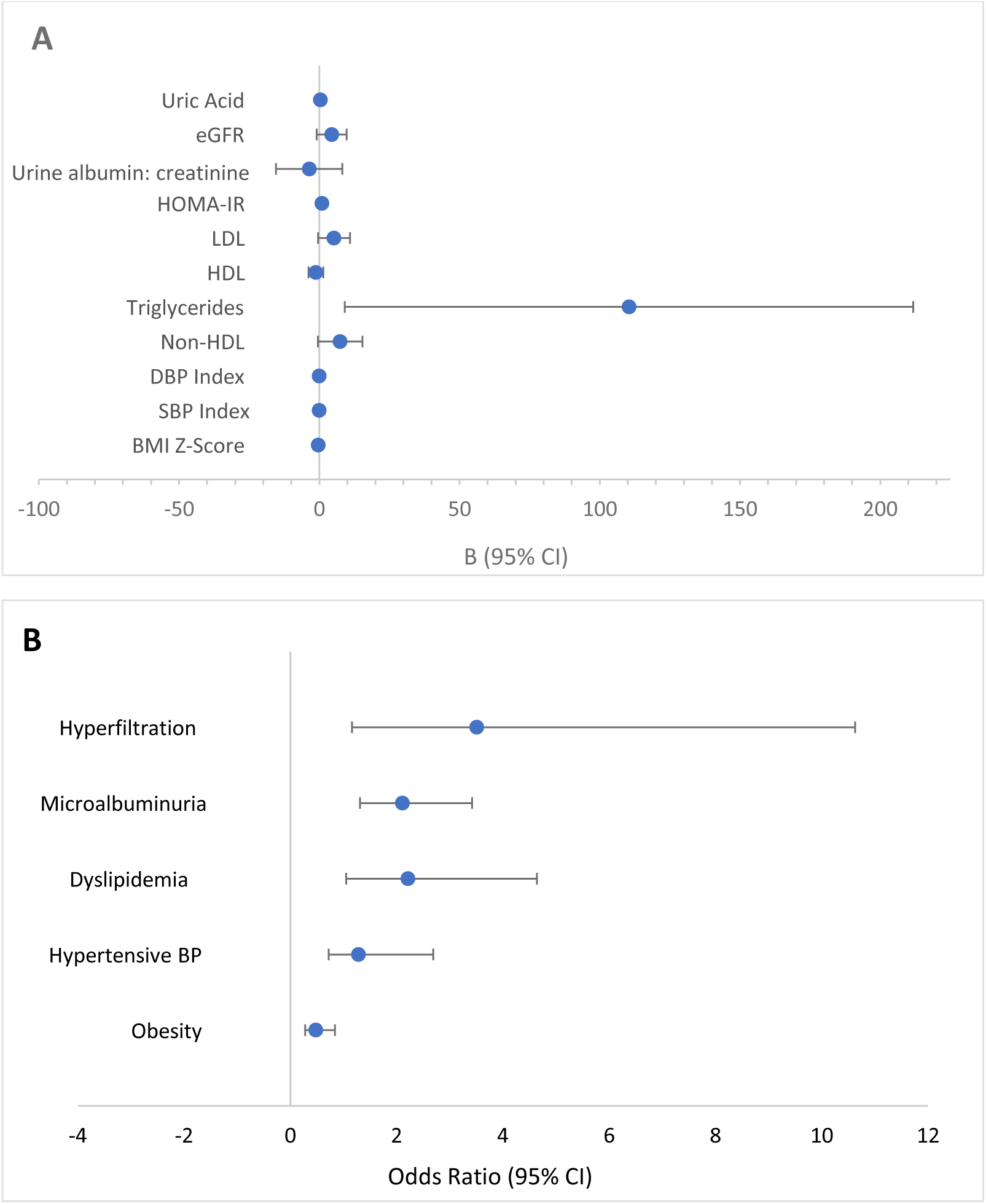
Forest plot displaying Asian American vs. Non Hispanic White (reference) cardiometabolic outcomes in adolescents. Models adjusted for age, sex, household income, food insecurity, passive smoke exposure and BMI z-score

### Cardiometabolic Variables: Foreign Born vs US Born

In adjusted regression analysis, there were no significant differences in obesity or blood pressure between US born and foreign born Asian American children. When compared to foreign born Asian American children, US born Asian American children had significantly higher non-LDL (b = 26.55, 95% CI 15.08, 38.03 p<0.001), triglycerides (b = 34.88, 95% CI 12.72, 57.04 p = 0.006), LDL (b = 19.82, 95% CI 5.85, 32.78 p = 0.01), HOMA-IR (b = 0.56, 95% CI 0.04, 31.09 p = 0.03), and uric acid (b = 0.63, 95% CI 0.38, 30.88 p<0.001) but also significantly lower HDL (b = -4.00, 95% CI -6.88, -1.13 p = 0.01) and eGFR (b = 11.97, 95% CI -18.90, -5.04 p = 0.004). US born Asian American children also had 0.0001 lower odds of hyperfiltration (Table 3 and Figure 2).

**Figure 2:**
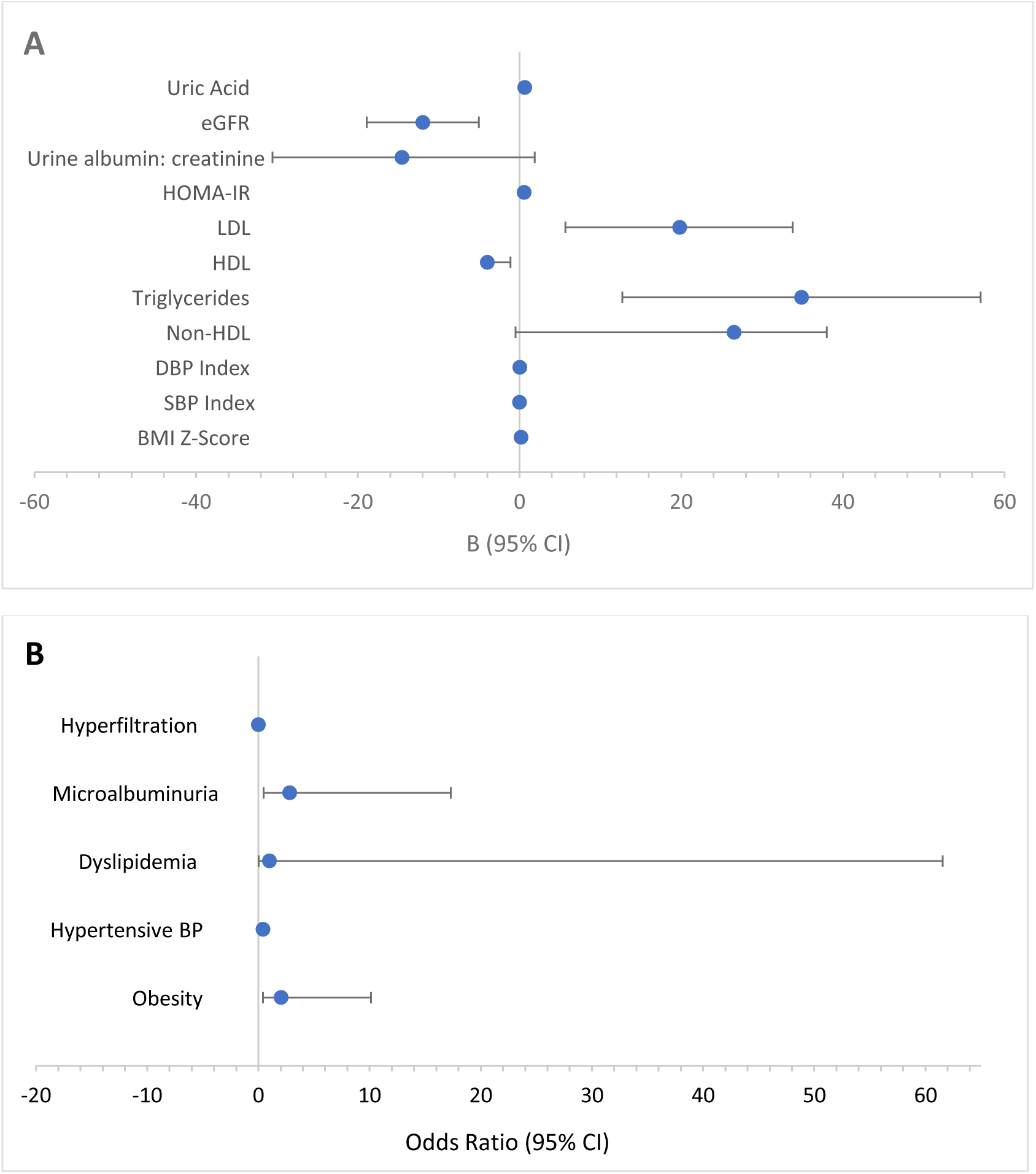
Forest plot displaying United States born vs foreign born (reference) cardiometabolic outcomes in Asian American adolescents. Models adjusted for age, sex, household income, food insecurity, passive smoke exposure and BMI z-score.

## Discussion

The aim of this study was twofold: to investigate cardiometabolic outcomes of Asian American children against those of NHWs and to investigate cardiometabolic health of US born and foreign born Asian American children. Adjusted models of the nationally representative sample showed that despite having lower BMI z-scores and lower odds of obesity, Asian American children had more insulin resistance and higher uric acid in addition to higher odds of dyslipidemia and impaired kidney function (microalbuminuria, and hyperfiltration) when compared to their NHW counterparts. Models also found that Asian American children born in the US had worse lipid markers, more insulin resistance, higher uric acid and lower eGFR when compared to foreign born Asian American children. No associations with blood pressure were found by racial group or place of birth in Asian American children.

Although our study found Asian American children to have significantly lower BMI z-score and lower odds of obesity compared to NHWs, higher BMI has historically been a poor predictor of certain diseases such as hypertension in Asian populations [14]. For example, Asian Americans have been documented in nationally representative samples presenting features of metabolic syndrome (e.g. obesity, high blood pressure, high blood triglycerides, low levels of HDL cholesterol, and insulin resistance) while at Western standards of normal BMI and weight [15]. For this reason, studies argue for the use Asian-specific BMI cut-points [14]. One study looking at adolescents in a New York City high school with excess weight found that despite having lower average BMIs, Asian Americans had significantly higher degrees insulin resistance and triglycerides than their non-Asian peers [16]. Additional research on adults in Northern California found that certain Asian American subgroups to have increased risk of dyslipidemia despite also having lower average BMIs [17]. These data support our findings where despite lower average BMIs, Asian Americans experience higher HOMA-IR levels and higher odds of dyslipidemia.

In terms of kidney health, our study found Asian Americans to have 2.1 higher odds of microalbuminuria and 3.5 higher odds of hyperfiltration compared to NHWs. Using NHANES 2011-2014, Kataoka-Yahiro et al. similarly found that Asian American adults are more likely to have albuminuria than NHWs [18]. However, this study also found that Asian Americans had a lower risk of mild to moderately reduced eGFR (< 60 ml/min/1.73m2) when compared to NHWs [18]. Furthermore, one study using NHANES 1999-2012 found adolescents experiencing glomerular hyperfiltration had a significantly higher HOMA-IR levels (3.52 vs. 3.01; P = 0.001), independent of BMI z-score, corroborating our results in NHANES 2011-2018 where Asian American adolescents had higher HOMA-IR levels and higher odds of hyperfiltration than NHWs [19].

With regard to acculturation, data is even more limited in children; however, studies in Asian American adults generally show increased acculturation correlated with poorer health [5,20–23]. Echeverria et al. studied adults using NHANES 2011-2014 and found that Asian Americans had an increased prevalence for CVD risk factors with longer durations in the US [5]. Analysis of NHANES 2011-2012 found that that foreign born Asian Americans were at elevated risk for high total cholesterol and LDL levels [24]. These results contrast with our findings of higher non-LDL (b=26.55, p<0.001), higher LDL levels (b=19.82, p=0.01), and lower HDL levels (b=-4.00, p=0.01) amongst US born Asian American adolescents. Additionally, Morey et al. found that longer residencies in the US was associated with higher triglyceride levels in Chinese and Korean American adults but failed to include other Asian subgroups and children in their sample [22]. The lack of uniformity in the literature and our findings further emphasize need for more work on the health implications of Asian American acculturation.

Several potential explanations may account for these associations between poor cardiometabolic outcomes, Asian American race, and acculturation. We hypothesized that acculturation to Western lifestyle in the US may involve the transition to a diet that is higher in fat, sugar, and processed foods which may lead to poorer health [25] . However, contrary to expectations, dietary intake was similar between Asian Americans and NHWs, with the exception of sugar consumption. No differences in dietary intake were found between Asian American children who were US born and foreign born. Because two-thirds of the Asian adolescents in our sample were US born, we speculate that their higher cardiometabolic risk was dually influenced by genetics and by epigenetic changes due to lifestyle influences. This dual influence provides more clarity on the racial disparity of Asian American adults for conditions such as gout which has doubled in prevalence from 2011 to 2018 [26]. Studies have found individuals with Asian heritages to be genetically disposed to elevated uric acid levels and when combined with Western diets rich in purines and sedentary lifestyles; thus, the risk of complications such as hyperuricemia, gout, and cardiovascular risk are further elevated in Asian Americans [27,28]. This finding in adult health is important given the age of our adolescent study sample, where Asian American adolescents are found to have higher levels of uric acid than NHWs (b=0.33, p=0.04); however, this finding does not explain why foreign born Asian American adolescents also have higher uric acid levels (b=0.63, p=0.01).

There are several limitations in our analyses. First, we are not able to establish causative conclusions between acculturation and cardiometabolic outcomes due to the cross-sectional nature of the NHANES dataset. In addition, NHANES data may not be complete due to over/under sampling of the Asian American population including omitting data from undocumented immigrants. Next, this study investigated the role of acculturation on Asian American adolescent health, but only included birthplace nativity in this analysis. Future studies should aim to investigate the role of other measures of acculturation including language proficiency, years in the US, socioeconomic status, and health literacy [21]. In addition, since we are studying the pediatric population, the parental acculturation status might be worth additional consideration. In addition, our study did not include Asian American subgroups in the analysis since NHANES data does not disaggregate the category Asian American into subgroups (e.g. Chinese, Filipino, Asian Indian, etc.). Studies that analyze cardiometabolic health by Asian subgroup, often show significant disparities such as increased blood pressure and increased odds of hypertension in certain Asian subgroups such as Filipino and South Asian adolescents [28,29] and adults [21,23,30–32]. Our lack of subgroup aggregation may play a role in why we did not find significant disparities in blood pressure and hypertension status in our study. However, the fact that our study still found significant results despite not carrying analysis by subgroup adds to the strength and significance in our findings [29,33,34]. The most important strength of our study is the source of our data from NHANES, reflective of a large, nationally representative, and non-clinical sampling of the US population.

This study demonstrated that Asian American pediatric populations may experience heightened cardiometabolic risk, especially for those who were US born. These results may encourage physicians to closely monitor the cardiometabolic health of their Asian American pediatric patients regardless of BMI levels and pay particular attention to their acculturation level. This study, in conjunction with previous literature, highlights the clinical importance of race-aware care and early detection of cardiometabolic risk among Asian American children, especially as the Asian American population continues to experience significant growth [2]. Early detection of cardiometabolic risk factors in childhood is critical for early therapeutic intervention, decreasing disease progression, and minimizing complications. Future studies should aim to increase the literature about Asian American and racial differences in cardiometabolic outcomes such as kidney function, cholesterol and adiposity levels, and insulin resistance as well as consider the role of a broader-defined acculturation on cardiometabolic function.

## Data Availability

All data produced in the present study are available upon reasonable request to the authors

https://www.cdc.gov/nchs/nhanes/index.htm

## Statements & Declarations

Funding: Dr. Kevin Meyers is the recipient of NIH grant funding from 1R01DK131091-01A1 and 1R01HL162912-01A1.

Competing Interests: All authors declare they have no competing financial or non-financial interests.

Author Contributions: All authors contributed to the study conception and design. Material preparation, data collection, and analysis were performed by all authors. This first draft of the manuscript was written by all authors. All authors read and approved the final manuscript.

## Acknowledgments

No acknowledgements

## Compliance with Ethical Standards

Conflict of Interest: The authors declare that they have no conflict of interests to disclose.

Ethical Approval: Not applicable.

Informed Consent: Not applicable.

Consent to Publish: Not applicable

## References

[1] Monte LM, Shin HB. 20.6 Million people in the US identify as Asian, Native Hawaiian or Pacific Islander. United States Census Bureau 2022. https://www.census.gov/library/stories/2022/05/aanhpi-population-diverse-geographically-dispersed.html

[2] Budiman A, Ruiz N. Asian Americans are the fastest-growing racial or ethnic group in the U.S. Pew Research Center 2021. https://pewrsr.ch/3tbjILO

[3] Fang J, Zhang Z, Ayala C, Thompson-Paul AM, Loustalot F. Cardiovascular health among non-Hispanic Asian Americans: NHANES, 2011–2016. J Am Heart Assoc 2019;8:e011324. 10.18865/ed.29.2.287.

[4] Satish P, Sadaf MI, Valero-Elizondo J, Grandhi GR, Yahya T, Zawahir H, et al. Heterogeneity in cardio-metabolic risk factors and atherosclerotic cardiovascular disease among Asian groups in the United States. Am J Prev Cardiol 2021;7:100219. 10.1016/j.ajpc.2021.100219.

[5] Echeverria SE, Mustafa M, Pentakota SR, Kim S, Hastings KG, Amadi C, et al. Social and clinically-relevant cardiovascular risk factors in Asian Americans adults: NHANES 2011–2014. Prev Med (Baltim) 2017;99:222–7.

[6] Centers for Disease Control and Prevention. NHANES survey methods and analytic guidelines. Retrieved May 2019;8:2019.

[7] Centers for Disease Control and Prevention. NCHS Ethics Review Board (ERB) Approval 2016.

[8] Johnson CL, Dohrmann SM, Burt VL, Mohadjer LK. National health and nutrition examination survey: sample design, 2011-2014. vol. 162. 2nd ed. National Center for Health Statistics. Vital Health Stat; 2014.

[9] Centers for Disease Control and Prevention. Defining Child BMI Categories 2023.

[10] Control C for D, Prevention. NHANES survey methods and analytic guidelines 2019;8:2019.

[11] Flynn JT, Kaelber DC, Baker-Smith CM, Blowey D, Carroll AE, Daniels SR, et al. Clinical practice guideline for screening and management of high blood pressure in children and adolescents. Pediatrics 2017;140.

[12] Rae-Ellen Kavey, Vivek Allada, Daniels SR. Expert panel on integrated guidelines for cardiovascular health and risk reduction in children and adolescents: summary report. Pediatrics 2011;128:S213–56.

[13] Chiarelli F, Verrotti A, Morgese G. Glomerular hyperfiltration increases the risk of developing microalbuminuria in diabetic children. Pediatric Nephrology 1995;9:154–8. 10.1007/BF00860729.

[14] Chait A, Den Hartigh LJ. Adipose tissue distribution, inflammation, and its metabolic consequences, including diabetes and cardiovascular disease. Front Cardiovasc Med 2020;7:22.

[15] Cheung EL, Bell CS, Samuel JP, Poffenbarger T, Redwine KM, Samuels JA. Race and obesity in adolescent hypertension. Pediatrics 2017;139.

[16] Elsamadony A, Yates KF, Sweat V, Yau PL, Mangone A, Joseph A, et al. Asian Adolescents with Excess Weight are at Higher Risk for Insulin Resistance than Non-Asian Peers. Obesity 2017;25:1974–9.

[17] Frank ATH, Zhao B, Jose PO, Azar KMJ, Fortmann SP, Palaniappan LP. Racial/ethnic differences in dyslipidemia patterns. Circulation 2014;129:570–9.

[18] Kataoka-Yahiro M, Davis J, Gandhi K, Rhee CM, Page V. Asian Americans & chronic kidney disease in a nationally representative cohort. BMC Nephrol 2019;20:1–10.

[19] Lee AM, Charlton JR, Carmody JB, Gurka MJ, DeBoer MD. Metabolic risk factors in nondiabetic adolescents with glomerular hyperfiltration. Nephrology Dialysis Transplantation 2017;32:1517– 24.

[20] Han Y-Y, Forno E, Celedón JC. Acculturation and asthma in Asian American adults. J Allergy Clin Immunol Pract 2022;10:2752–2753. e1.

[21] Min LY, Islam RB, Gandrakota N, Shah MK. The social determinants of health associated with cardiometabolic diseases among Asian American subgroups: a systematic review. BMC Health Serv Res 2022;22:1–16.

[22] Morey BN, Ryu S, Shi Y, Park HW, Lee S. Acculturation and Cardiometabolic Abnormalities Among Chinese and Korean Americans. J Racial Ethn Health Disparities 2022:1–11.

[23] Shah MK, Gandrakota N, Gujral UP, Islam N, Narayan KMV, Ali MK. Cardiometabolic Risk in Asian Americans by Social Determinants of Health: Serial Cross-sectional Analyses of the NHIS, 1999– 2003 to 2014–2018. J Gen Intern Med 2023;38:571–81.

[24] Jackson HL. Risk of Type 2 Diabetes among US and Foreign Born Non-Hispanic Asians: Evidence from NHANES 2011-12 (2015) J Diabetes Obesity 2 (1): 5-8. J Diabetes Obes 2015;2.

[25] Lee JR, Maruthur NM, Yeh H-C. Nativity and prevalence of cardiometabolic diseases among US Asian immigrants. J Diabetes Complications 2020;34:107679.

[26] Yokose C, McCormick N, Lu N, Tanikella S, Lin K, Joshi AD, et al. Trends in Prevalence of Gout Among US Asian Adults, 2011-2018. JAMA Netw Open 2023;6:e239501–e239501.

[27] Butler F, Alghubayshi A, Roman Y. The epidemiology and genetics of hyperuricemia and gout across major racial groups: a literature review and population genetics secondary database analysis. J Pers Med 2021;11:231.

[28] Vuskan V, Rogovik A, Jenkins A, Peeva V, Beljan-Zdravkovic, Stavro M, et al. Cardiovascular risk factors, diet, and lifestyle among European, South Asian and Chinese adolescents in Canada. Paediatr Child Health 2012;17:e1–6.

[29] Cook WK, Tseng W, Bautista R, John I. Ethnicity, socioeconomic status, and overweight in Asian American adolescents. Prev Med Rep 2016;4:233–7.

[30] Li Y, Alicia ZHU, Austin LE, Singh J, Palaniappan LP, Srinivasan M, et al. Association of Acculturation with Cardiovascular Risk Factors in Asian-American Subgroups. Am J Prev Cardiol 2023;13:100437.

[31] Kizzee OP, Lo JC, Ramalingam ND, Rana JS, Gordon NP. Differential cardiometabolic risk factor clustering across US Asian ethnic groups. Am J Prev Med 2022;62:e129–31.

[32] Jih J, Mukherjea A, Vittinghoff E, Nguyen TT, Tsoh JY, Fukuoka Y, et al. Using appropriate body mass index cut points for overweight and obesity among Asian Americans. Prev Med (Baltim) 2014;65:1–6.

[33] Jacob M, Cho L. Asian Americans and cardiometabolic risk: why and how to study them. J Am Coll Cardiol 2010;55:974–5.

[34] Palaniappan LP, Araneta MRG, Assimes TL, Barrett-Connor EL, Carnethon MR, Criqui MH, et al. Call to action: cardiovascular disease in Asian Americans: a science advisory from the American Heart Association. Circulation 2010;122:1242–52.

